# Next-Generation Phenotyping: Introducing PhecodeX for Enhanced Discovery Research in Medical Phenomics

**DOI:** 10.1101/2023.06.18.23291088

**Authors:** MM Shuey, WW Stead, I Aka, AL Barnado, JA Bastarache, E Brokamp, MS Campbell Joseph, RJ Carroll, JA Goldstein, A Lewis, BA Malow, JD Mosley, T Osterman, DA Padovani-Claudio, A Ramirez, DM Roden, BA Schuler, E Siew, J Sucre, I Thomsen, RJ Tinker, S Van Driest, C Walsh, JL Warner, QS Wells, L Wheless, L Bastarache

## Abstract

**Summary:** Phecodes are widely-used and easily adapted phenotypes based on International Classification of Diseases (ICD) codes. The current version of phecodes (v1.2) was designed primarily to study common/complex diseases diagnosed in adults. Here we present phecodeX, an expanded version of phecodes with a revised structure and 1,761 new codes. PhecodeX adds granularity to phenotypes in key disease domains that are under-represented in the current phecode structure-including infectious disease, pregnancy, congenital anomalies, and neonatology- and is a more robust representation of the medical phenome for global use in discovery research.

**Availability and implementation:** phecodeX is available at https://github.com/PheWAS/phecodeX.

**Supplementary information:** Supplemental Tables 1-4, Bastarache_bioRxiv_20220907.pdf

WC-1999

## Introduction

Phecodes are manually curated groups of International Classification of Diseases (ICD) codes intended to capture clinically meaningful concepts for research (Denny, et al., 2010). Although initially created to conduct phenome-wide association studies (PheWAS) of common genetic loci, the applications of phecodes have broadened considerably in recent years (Bastarache, 2021). Phecodes have been used in conjunction with various methods – including machine learning, comorbidity clustering, and prediction algorithms – and to conduct research across a wide array of clinical domains, from Mendelian disease to pharmacology (Bastarache, et al., 2022; Hellwege, et al., 2023; McArthur, et al., 2023; Pruett, et al., 2021; Zhang, et al., 2021).

Because they are based on ICD codes – a global standard for classifying disease – phecodes can be used in virtually every electronic health record (EHR)-linked biobank, both in the United States and internationally (Zhou, et al., 2022). The wide-spread use of phecodes attests to their value, however, the structure has not been revised since 2013, and the current version (v1.2) has limitations. Phecode v1.2 focused on capturing diseases present in the genome-wide association (GWAS) catalog, which was comprised mainly of common diseases of adulthood (Denny, et al., 2013; Sollis, et al., 2023). Phecodes relating to pregnancy, congenital anomalies, and neonatology are present in v1.2, but only as highly aggregated concepts. Furthermore, v1.2 was designed using the outdated ICD-9 coding system and does not take advantage of the greater granularity and modernized organization of ICD-10s. Here we present phecodeX, an expanded and updated version of phecodes designed to overcome many of these limitations and further facilitate new and creative phecode applications.

### phecodeX

PhecodeX was created via manual curation in consultation with 22 clinicians and domain experts. Each expert received a draft version of phecodeX according to their expertise and provided feedback. Changes were discussed with iterative adjustments made until a final grouping was determined. This method was also employed to create phecode v1.2, although the collective expertise of clinical domain experts was greater for phecodeX. The final mapping schema is available for download (https://github.com/PheWAS/phecodeX) and summarized in Supplemental Tables 1-2 (includes mapping files that support ICD-10 and ICD-10-CM (clinical modification)).

The new phecodeX map differs from v1.2 in several ways. PhecodeX 1) *aligns its structure with the ICD-10 coding system*, 2) *revises the phecode labeling system*, 3) *leverages multi-mapping of both ICD-9 and -10 codes*, 4) *removes exclude ranges used to define controls*, and 5) *reorganizes phecode categories*. Because of these changes, which are detailed below, phecodeX is a more comprehensive representation of the medical phenome with enhanced coverage of phenotypes relevant to both complex and monogenic disease (Figure 1).

**Figure 1.**
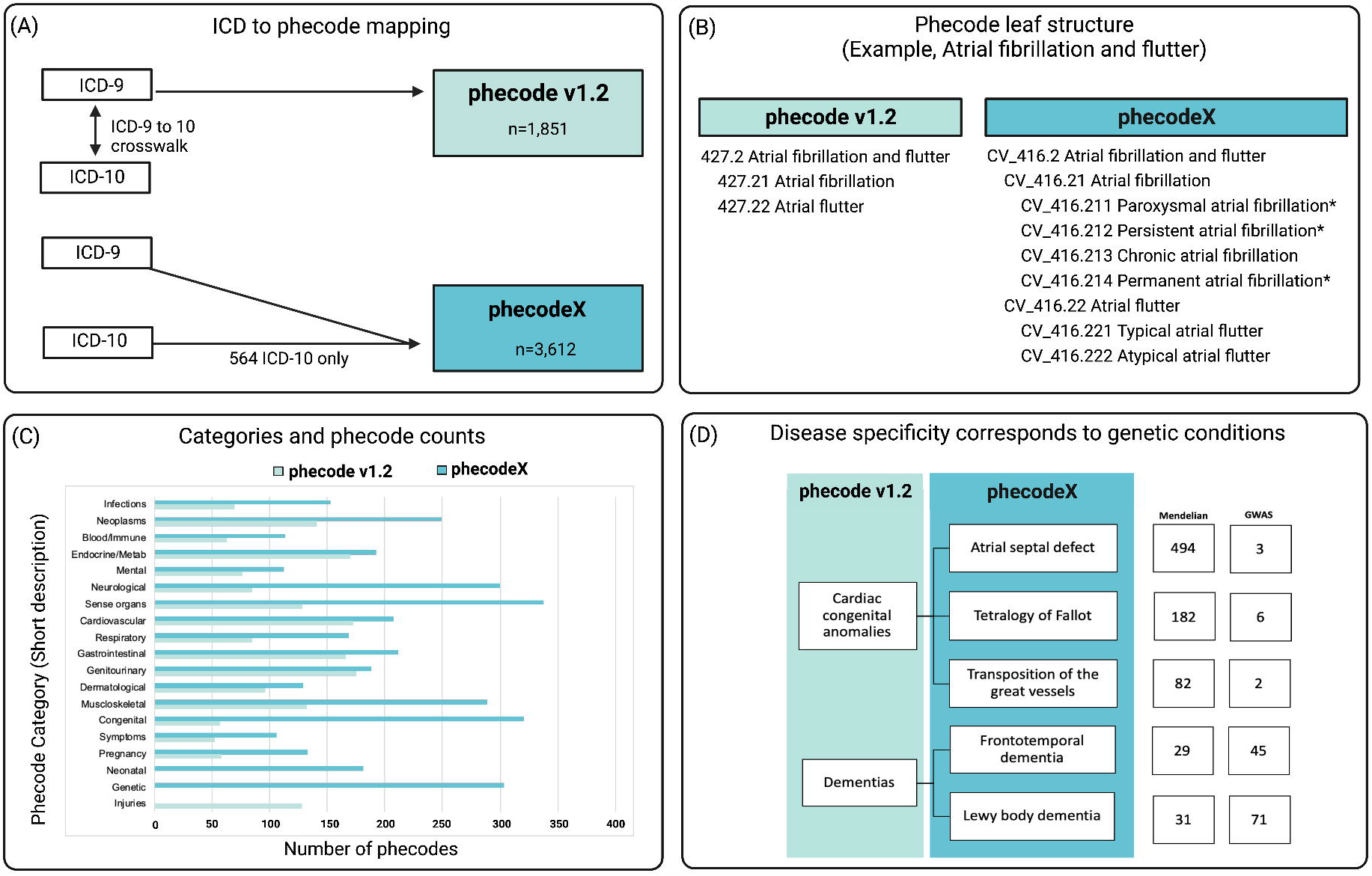
Overview of the phecodeX data structure and differences from version 1.2. Panel (A) demonstrates how phecode v1.2 was designed using codes from the International Classifiers of Disease, ninth revision (ICD-9); ICD-10s were later integrated into phecodes using the ICD crosswalk. PhecodeX adapted the overall structure of ICD-10s; codes from ICD-9 and -10 were mapped simultaneously to optimize the phecode structure for both ICD versions. Panel (B) provides an example of the expansion in phecode tree structure. The new system adds a tertiary level with up to three digits past the decimal place for increased phecode specificity. This example also demonstrates the difference in phecode labels and strings, including the introduction of a two-character category descriptor (e.g., CV for the Cardiovascular category) and the use of * after code description to denote codes that map to only ICD-10 billing codes. Panel (C) demonstrates the difference in the size of phecodeX versus v1.2, stratified by phecode category. Finally, panel (D) shows how new phecodes introduced in phecodeX improve coverage of phenotypes relevant to both complex and Mendelian disease. In this example, the two phecodes in v1.2 are expanded to reflect five phecodes in phecodeX. The column labeled “Mendelian” indicates the number of Mendelian disease genes linked to the phenotypes through the Human Phenotype Ontology (HPO). The column labeled “GWAS” indicates the number of unique genetic variants present in the Genome-wide association study (GWAS) catalog. Figure was generated using BioRender.

### Alignment with ICD-10 structure

By updating its structure to *align with the ICD-10 coding system*, phecodeX takes advantage of the increased granularity available in the ICD-10 schema. The ICD-10 coding structure includes nearly ten times as many codes compared with ICD-9(Fung, et al., 2016; Steindel, 2010). While ICD-10 codes have been integrated into v1.2, no new phecodes were introduced in this process(Wu, et al., 2019). Thus, the design of v1.2 was based on ICD-9 and did not take advantage of the increased specificity of ICD-10s. PhecodeX includes 572 new phecodes (Figure 1A) representing diagnoses that were newly added to the ICD-10 system (e.g., Dilated cardiomyopathy, COVID-19, and Gestational diabetes). Phecodes supported by ICD-10 alone are denoted in the icd10_only column of the mapping file and their phecode string ends with an “*” (Supplemental Table 1).

PhecodeX also reflects a more modern organization of disease classification. ICD-10s became mandatory in 2015 in the United States and are structured based on current understandings of disease etiology, which evolved since the introduction of ICD-9s in 1977(Fung, et al., 2016). For example, the ICD for Macroglobulinemia was classified in the “Endocrine/Metabolic” chapter in ICD-9s and the “Neoplasm” chapter in ICD-10s. These changes in disease classification are reflected in the phecodeX coding structure.

### New phecode labels

Each phecodeX label is prefixed by a two-letter label indicating the category, followed by an underscore and a three-digit root code. In contrast, v1.2 phecodes were labeled with three-digit root codes, similar to ICD-9s. The character prefixes make phecodeX visually distinct from v1.2 and ICD codes, and prevent programs like R and Excel from corrupting codes by interpreting them as integers (e.g., phecode “008” being transformed to “8”). The numeric component of each phecodeX code is unique, even without the prefix.

PhecodeX allows for three decimals to indicate child codes, as opposed to v1.2 that allowed for a maximum of two digits. By adding a tertiary level of specificity, phecodeX includes more granular codes than were possible in v1.2. For example, atrial fibrillation and flutter form the basis for the code CV_416.2 (427.2 in v1.2) which is further delineated into two secondary child codes -- fibrillation (416.21) and flutter (416.22) – which are the final specification for these conditions. In phecodeX, a tertiary child provides additional granularity reflecting greater specificity, e.g., atrial flutter (CV_416.22) is further separated into typical and atypical flutter (CV_416.221 and CV_416.222, respectively) (Figure 1B).

### Multi-mapping

PhecodeX embraces multi-mapping where one ICD code can map to more than one phecode. In contrast, v1.2, is based on a 1-to-1 mapping (each ICD-9 mapped to a unique phecode). The 1-to-1 constraint of v1.2 led to difficulties in classifying combination ICD codes (i.e., codes that indicate multiple diagnoses), particularly for infectious disease (ID) diagnoses which were often indicated by combination ICDs in body-specific chapters (Lu, 2005). For example, in v1.2 the ICD for “Pneumonia due to staphylococcus” is mapped to 480.1 (*Bacterial pneumonia*) but not 041.1 (*Staphylococcus infections*), while in phecodeX, this ICD is mapped to both phenotypes (ID_009 and RE_468.2, respectively).

Multi-mapping enabled a full restructuring of the phecodeX ID category, whereby all infectious agents are organized by genus for bacteria and fungi, and family for viruses. ICDs from the perinatal chapters are also frequently multi-mapped, such that the phecode for *Type 1 diabetes* (EM_202.1) includes ICDs from the endocrine/metabolic chapter (E10*), as well as perinatal codes like O24.01 (*Pre-existing type 1 diabetes mellitus, in pregnancy*).

### Removal of excluded ranges

PhecodeX does not include specific exclude ranges for the controls. Exclude ranges are lists of phecodes that are used to remove controls with potentially related conditions. Phecode v1.2 includes an exclude range for each phecode, and they are implemented by default in the PheWAS R package (Carroll, et al., 2014), However, recent research suggests that exclude ranges do not globally improve the ability for phecodes to replicate known genetic associations (Bastarache, et al., 2022). Due to the complexity introduced by exclude ranges, both in terms of executing and interpreting PheWAS analyses, combined with limited evidence for their benefit, we elected not to include these ranges in phecodeX.

### phecodeX Categories

Phecodes are grouped into categories, similar to ICD chapters. PhecodeX includes 18 categories that largely mirror those in v1.2 with three exceptions (Figure 1C). First, phecodeX has a category for neonatal conditions. In v1.2, both neonatal and pregnancy-related phecodes fall under the category of “pregnancy complications.” Second, phecodeX eliminates the v1.2 category for “injuries & poisonings,” which includes 128 phecodes for traumatic injuries, poisonings, and surgical complications. In phecodeX, codes for fractures and dislocations have been moved to the musculoskeletal category. The remaining “injuries & poisonings” codes are excluded from phecodeX. While these codes may prove highly valuable for research, they require specific exposures to manifest and thus are ill-suited to a PheWAS-style analysis. Finally, phecodeX introduces a new section for genetic conditions that includes 324 phecodes for specific genetic diagnoses and chromosomal anomalies (e.g., Rett syndrome, Trisomy 18, and DiGeorge syndrome). Phecodes in the genetic category are defined by ICD codes that indicate a disease that is solely attributed to a genetic defect; diseases that have both genetic and non-genetic causes are not included. A summary of phecodeX by category can be found in Supplemental Table 3.

### phecodeX and multiple testing burden

Due to these changes, phecodeX has nearly twice the number of phecodes as v1.2 (3,612 vs. 1,851, respectively). PhecodeX expanded granularity is most notable in categories for pregnancy, neonatal, and congenital anomalies. PhecodeX includes 5.8 times more codes for congenital anomalies compared with v1.2 (365 vs 56), and 5.1 times more codes for pregnancy/neonatal disorders (297 vs. 58).

The expansion in granularity has the benefit of a more comprehensive representation of the medical phenome, including many conditions associated with rare and common genetic drivers of disease (Figure 1D). However, the addition of new codes can also impact multiple testing burden for analyses such as PheWAS. If all codes were included in a PheWAS, the Bonferroni corrected P value for nominal significance using v1.2 would be 2.70 × 10^−5^ versus 1.38 × 10^−5^ for phecodeX. However, this difference is mitigated by the fact that many new codes in phecodeX are for rare phenotypes and diagnoses. While these codes are important for studying rare genetic diseases, many will be excluded from PheWAS analysis due to low case counts. For example, in a cohort of 72,024 individuals drawn from BioVU – Vanderbilt’s de-identified biobank – the number of phecodes with at least 100 cases was 1,392 for v1.2 and 1,583 for phecodeX (Bonferroni P of 3.59 × 10^−5^ versus 3.16 × 10^−5^).

The testing burden can further be alleviated by removing or restricting to categories relating to relevant life stages, a major improvement in the new version. For example, if a researcher is interested in identifying pregnancy-related complications phecodeX would allow for a more restrictive analysis focusing on the relevant category. This is because the newer version allows for multi-mapping that would allow for conditions relating to pregnancy, such as pregnancy-related diabetes, to map to both the endocrine and the pregnancy-related conditions categories.

### Phecode v1.2 to phecodeX crosswalk

We created a crosswalk between v1.2 and phecodeX. In total, 1,055 of the 1,851 (57%) phecodes in v1.2 map to codes in phecodeX (Supplemental Table 4). There are a number of reasons that the remaining 43% of v1.2 codes are not present in phecodeX. First, in designing phecodeX, we avoided creating vague or non-specific codes, insofar as it was possible given the constraints of the ICD structure. In v1.2, 213 codes (12% of all phecodes) include the string “Other”, “NOS” (not otherwise specified), or “NEC” (not elsewhere classified). While these codes may contain valuable phenotypic information, their vague labels make interpretation difficult. In phecodeX, the number of non-specific phecodes was reduced to 91 (2.0% of all phecodes). Second, because the two versions of phecodes were designed based on different ICD coding systems (ICD-9 versus ICD-10), parent phecodes group multiple conditions under different parent codes.

The majority of common/complex diseases present in v1.2 are present in phecodeX and reflect the intent of v1.2 to capture diseases present in the GWAS catalog. Of the 149 phecodes in v1.2 that map to GWAS catalog diseases, 138 (93%) are present in phecodeX. Therefore, PheWAS conducted on common genetic variants may not be significantly different between the two versions. However, analysts should be aware that phecodes represented in both versions may have underlying differences in terms of their ICD groupings and the improvements in diagnostic specificities in the latter may improve the capture of particular physiologic conditions.

### Compatibility with existing software packages

Additional supplemental files for data use and comparison with v1.2 as well as example code that will facilitate the use of phecodeX with the PheWAS package is available at https://github.com/PheWAS/phecodeX.

## Conclusions

PhecodeX (version 1.0) represents the first major revision of phecodes since the release of v1.2 in 2013. The increased specificity of phecodeX provides more granular coverage of the medical phenome intended to support new uses for phecodes and encourages further creative applications. Indeed, an alpha version of phecodeX has already been used in numerous publications, including studies on pregnancy and neonatal outcomes and hereditary cancer syndromes – studies that would not have been feasible without the additional granularity offered in phecodeX (Campbell, et al., 2023; Stead, et al., 2023; Zeng, et al., 2022). We acknowledge that v1.2 remains suitable for large-scale PheWAS, particularly for studies focused on common/complex diseases. Furthermore, many existing catalogs use v1.2, therefore, researchers may want to continue to use v1.2 for cross-site meta-analyses or replications until these resources adopt phecodeX (Zawistowski, et al., 2023). Due to the broader and more in-depth coverage of the clinical ICD-based phenome and early adaptation of the method, we anticipate phecodeX will be a broadly applied resource for medical informatics.

Future iterations of phecodeX may integrate suggestions from the broader research community to enhance specificity and applicability.

## Data Availability

The presented software is available at: https://github.com/PheWAS/phecodeX.

